# LDCT-based Lung Cancer Screening and Small Cell Lung Cancer: limited but non-negligible impact on survival. A brief report

**DOI:** 10.1101/2025.06.09.25329254

**Authors:** Roberta Eufrasia Ledda, Federica Sabia, Camilla Valsecchi, Luigi Rolli, Margherita Ruggirello, Alfonso Vittorio Marchianò, Ugo Pastorino

## Abstract

**Introduction:** Low-dose computed tomography (LDCT)-based lung cancer screening (LCS) seems to have very limited impact on small cell lung cancer (SCLC) outcomes. This study aims at describing frequency and outcomes of SCLC in a large LCS population.

**Methods:** Patients with a histological diagnosis of SCLC among the participants of 3 different trials (7473) were selected for the analysis. Demographic and clinical data were collected at the baseline and follow-up screening rounds, while the vital status and date of death were obtained through a dedicated national platform.

**Results:** Of the 396 diagnosed LCs, 28 (7.1%) were SCLCs; median survival time from the diagnosis was 1.5 years, and overall mortality was 71.4%. Screen-detected SCLCs were 20/28 (71.4%); 5/20 (25%) were prevalent cancers and 15/20 (75%) incident ones. Five-year mortality among the screen-detected and non-screen detected SCLCs was 70% and 62.5%, respectively.

**Conclusion:** The frequency of SCLC was lower as compared to other trials. Although no significant differences in 5-year mortality were observed between screen-detected and non-screen-detected SCLCs, the overall 5-year mortality was substantially lower than that reported in non-LCS populations, suggesting that LDCT-based LCS has an impact on SCLC outcome, albeit limited.

## Introduction

Small cell lung cancer (SCLC) is a neuroendocrine carcinoma accounting for approximately 15% of all cases of lung cancer (LC), characterized by rapid growth and early widespread metastases, with only one third of patients presenting with limited disease at the time of diagnosis and a 5-year mortality exceeding 90% [1, 2]. Nearly all cases of SCLC are attributable to cigarette smoking and the 2-3% of cases diagnosed in never-smokers are thought to be secondary to environmental and/or occupational exposures (e.g., radon) [1].

Although low dose computed tomography (LDCT) – based lung cancer screening (LCS) allows for detection of early-stage non-small cell lung cancer (NSCLC), it does not seem to be effective for detecting early-stage SCLC [3]. This is likely due to the aggressiveness of such a type of cancer that becomes symptomatic between consecutive screening rounds, thereby limiting the potential reduction of LC-related mortality [2]. Indeed, previous literature demonstrated not impact of LDCT-LCS on SCLC outcomes [4, 5].

This study aims at describing the frequency and the outcome of SCLC in a large LCS population.

## Materials and Methods

Data were retrospectively collected from three LDCT LCS trials conducted at the Fondazione IRCCS Istituto Nazionale dei Tumori of Milan from September 2005 to March 2020. Detailed information on these trials have been provided elsewhere [6-8]. Briefly, from 2005 to 2011 the Multicentric Italian Lung Detection (MILD) trial (ClinicalTrials.gov Identifier: NCT02837809) enrolled 4099 volunteers: 1723 were randomized to the control group and 2376 to the LDCT arm. All subjects included in the latter group were considered potentially eligible in the present analysis. The bioMILD trial (clinicaltrials.gov ID: NCT02247453) enrolled 4119 volunteers between 2013 and 2016 combining LDCT with plasma microRNA (miRNA) for personalised screening intervals. The Screening and Multiple Intervention on Lung Epidemics (SMILE, clinicaltrials.gov ID: NCT03654105) study enrolled 978 volunteers from 2019 to 2020 and offered LDCT screening in combination with a multifactorial preventive intervention (i.e., smoking cessation therapy with cytisine and reduction of chronic inflammation with low-dose acetylsalicylic acid in heavy tobacco users).

The original Institutional Review Board approval and written informed consent allowed the use of data for future research, including this analysis.

All patients with a histological diagnosis of SCLC among the 7473 screened subjects were selected for the present analysis. Demographic and clinical data were collected at the baseline and follow-up screening rounds, including age, sex, smoking status, pack-years and stage of disease (according to the 8^th^ edition TNM stage classification [9]). The vital status and date of death were obtained through the Istituto Nazionale di Statistica (ISTAT, SIATEL 2.0 platform). Participants accumulated person-years of follow-up from the baseline LDCT until death or the last available follow-up as of June 2024. Continuous variables were presented as median values and interquartile ranges (IQR), and categorical variables were reported as numbers and percentages. Survival curves were estimated by the Kaplan-Meier method.

## Results

A total of 7473 screened volunteers accumulated 82×1000 person-years. Of the 396 LCs diagnosed in 355 volunteers, 28 (7.1%) were SCLCs. SCLCs were diagnosed in 12 (42.9%) female and 16 (57.1%) male participants, median age at diagnosis was 67.5 years (Table 1). At the baseline round, 25/28 (89.3%) were current smokers, with median pack-years of 64.5. Three out of 28 (10.7%) were stage I, 3 (10.7%) stage II, 11 (39.3%) stage III and 11 (39.3%) stage IV. Only 8 SCLC were surgically resected. Median survival time from the diagnosis was 1.5 years, and overall mortality was 71.4% (20/28). Thirty-days mortality was 0, 90-days mortality 3.6%, 1-year mortality 21.4% and 5-year mortality 67.9% (19/28). Six patients (21.4%) were still alive at 5-year follow-up and 2 at >8-year (Figure 1). Screen-detected SCLCs were 20/28 (71.4%), with a median time of diagnosis from baseline of 3.8 years; 5/20 (25%) were prevalent cancers and 15/20 (75%) incident ones. Among the incident SCLCs, 1 (6.7%) was stage I, 1 (6.7%) stage II, 6 (40%) stage III, and 7 (46.6%) stage IV. Five-year mortality was 70% (14/20) and 3/20 (15%) patients, who underwent chemo- and radiotherapy, had a survival time >5 years from diagnosis and were still alive at the last follow-up (1 stage II, 1 stage III, 1 stage IV) (Table 2).

**Table 1.**
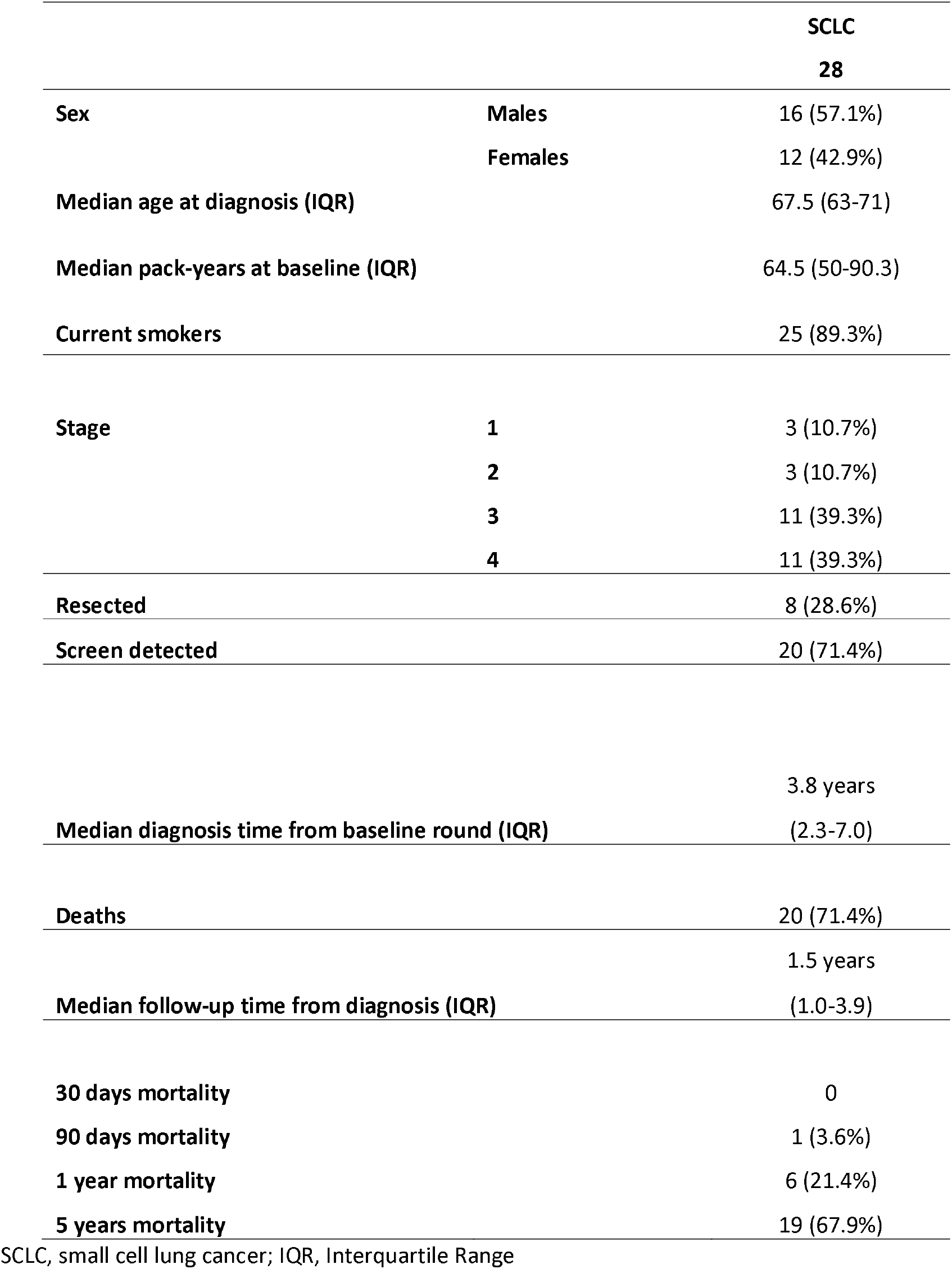
Demographics, clinical characteristics and outcome of screenees diagnosed with small cell lung cancer.

**Table 2.** Clinical characteristics and outcomes of screenees with screen-detected small cell lung cancer.

|  |  | screen detected SCLC |
| --- | --- | --- |
|  |  | 20 |
| **Stage** | **1** | 1 (5%) |
|  | **2** | 2 (10%) |
|  | **3** | 8 (40%) |
|  | **4** | 9 (45%) |
| **Resected** |  | 5 (25%) |
| **Median diagnosis time from baseline round (IQR)** |  | 3.1 years<br>(1.8-5.4) |
| **Deaths** |  | 14 (70%) |
| **Median follow-up time from diagnosis (IQR)** |  | 1.4 years<br>(0.8-2.3) |
| **30 days mortality** |  | 0 |
| **90 days mortality** |  | 1 (5%) |
| **1 year mortality** |  | 5 (25%) |
| **5 years mortality** |  | 14 (70%) |

**Figure 1.**
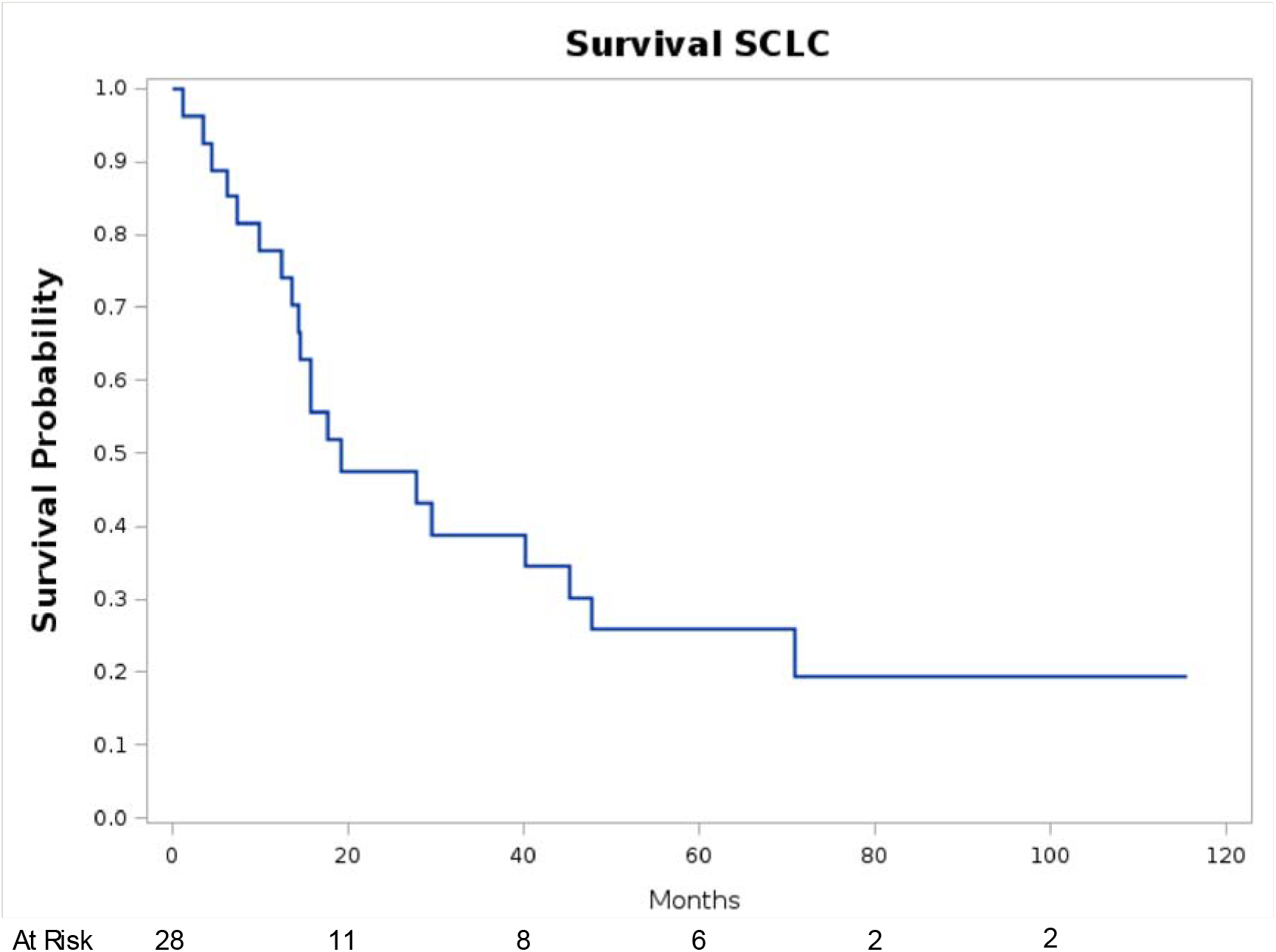
Overall survival of screenees diagnosed with small cell lung cancer.

Five out of 8 non-screen-detected SCLC were diagnosed after the end of the screening program (4 over 7 years from the baseline round), while the other 3 SCLC were interval detected. Five-year mortality for the non-screen detected SCLCs was 62.5% (5/8).

## Discussion

We observed a SCLC frequency of 7.1% with a slightly higher prevalence in male participants. Most SCLCs (71.4%) were screen-detected and among these, 75% were depicted at the incident round. The overall frequency was lower as compared to other trials, but similarly to previous reports most SCLCs (78.6%) were diagnosed at either stage III or IV [4, 10, 11].

Overall median survival time was 1.5 years, while 5-year mortality was 71.4%. In line with previous studies, no significant differences in 5-year mortality were observed between screen-detected and non-screen-detected SCLCs. Nevertheless, the overall 5-year mortality was substantially lower than that reported in non-LCS populations (>90%) [1]. It can be speculated that the smoking cessation support that was offered to the current smokers might have played a role in reducing the mortality rate. Moreover, only 3 of the 8 non-screen-detected SCLCs were interval detected cancers, whereas 5 were depicted at the end of the programme, suggesting that the continuations of LCS in older adults (e.g., up to 80 years of age) might have a positive impact on both NSCLC and SCLC [12].

Having said that, our results confirm that the impact of LDCT-LCS on SCLC outcomes remains limited. Recent research demonstrated that tumor DNA and other cancer-derived components detectable in the bloodstream allow for identification of tumor-specific mutations, suggesting that liquid biopsy may complement imaging modalities, especially in high-risk subjects in whom LDCT alone is not sufficient [13].

This study has some limitations. First, as inherent to the nature of any retrospective analysis, our observations are subject to confounding factors. Second, the small number of SCLC cases limits the possibility of comparison between screen-detected and non-screen-detected SCLC and between SCLC and NSCLC. Nonetheless, the LCS duration and length of follow-up allows for an assessment of long-term outcomes.

In conclusion, the SCLC-related mortality seems to be lower in the LCS population than in the general population, suggesting that the impact of LDCT-based LCS on SCLC is non-negligible, albeit limited.

## Data Availability

All data produced in the present study are available upon reasonable request to the authors

